# Optimising Early Management of Acute Severe Ulcerative Colitis in the Biologics Era: Admission Model for Intensification of Therapy in Acute Severe Colitis (ADMIT–ASC)

**DOI:** 10.1101/2021.07.14.21260477

**Authors:** Alex Adams, Vipin Gupta, Waled Mohsen, Thomas P Chapman, Deloshaan Subhaharan, Pradeep Kakkadasam Ramaswamy, Sudheer Kumar Vuyyuru, Saurabh Kedia, Colleen GC McGregor, Tim Ambrose, Bruce D George, Rebecca Palmer, Oliver Brain, Alissa Walsh, Vineet Ahuja, Simon Travis, Jack Satsangi

**Affiliations:** Translational Gastroenterology Unit, NIHR Oxford Biomedical Research Centre, Oxford University Hospitals NHS Foundation Trust, Oxford, UK; Department Of Gastroenterology, Cardiff University Health Board, UK; Digestive Diseases Unit, Gold Coast University Hospital, Queensland, Australia; Department of Gastroenterology, Western Sussex NHS Foundation Trust; Department of Gastroenterology, All India Institute of Medical Sciences (AIIMS), New Delhi, India

**Author notes:** Corresponding: John Radcliffe Hospital (Level 5), Headley Way, Headington, Oxford, OX3 9DU., /. Joint first author (Alex Adams, Vipin Gupta, Waled Mohsen).

**Keywords:** Acute severe colitis, prediction, outcome

## Abstract

**Background & aims:** We aimed to determine whether changes in ulcerative colitis management have translated to improved outcomes, in order to develop a simple model to predict steroid non-response on admission.

**Methods:** Outcomes of 131 adult ASC admissions (117 patients) in Oxford, UK between 2015-19 were compared with prospectively collected data from 1992-3. All patients received standard treatment with intravenous corticosteroids and endoscopic disease activity scoring (UCEIS). Steroid non-response was defined as receiving rescue medical therapy or surgery. A predictive model created in the Oxford cohort was validated in Australia and India (110 hospitalised patients Gold Coast University Hospital 2015-20; 62 hospitalised patients AIIMS, New Delhi 2018-20).

**Results:** In the 2015-19 Oxford cohort, 71 (54%) patients received medical rescue therapy (27% ciclosporin, 27% anti-TNF), compared to 27% ciclosporin in 1992-3, p=0.0015. Only 15% required colectomy during admission vs 29% in 1992-3 (p=0.033). Admission CRP, albumin, and UCEIS scores predicted steroid non-response (FDR p=0.00066, 0.0066 and 0.015). A four-point model was developed involving CRP ≥100mg/L (1 point), albumin ≤25g/L (1 point), UCEIS ≥4 (1 point) or ≥7 (2 points). Scoring 0 or 4 was 100% predictive of steroid response and non-response, respectively, in all three cohorts. Patients scoring 3-4 had 83% risk of steroid non-response in Oxford and 84% (0.70-0.98) in the validation cohorts – OR 11.9 (10.8-13).

**Conclusion:** Colectomy rates for ASC have halved in 25 years, while use of rescue medical therapy has doubled. Patients who are highly unlikely to respond to parenteral steroid treatment alone may be readily identified on admission, to be prioritised for early intensification of therapy.

**WHAT YOU NEED TO KNOW:** *Background and context:* Acute severe colitis (ASC) requires hospitalisation and, frequently, colectomy. The accepted management of ASC is three-five days of intravenous steroids followed by protocolised response assessment.

*New findings:* Medical rescue therapy has doubled and the colectomy rate halved in 25 years. A predictive index identifying steroid non-response was derived and validated in three independent centres in three countries.

*Limitations:* A need remains to demonstrate that earlier rescue medical therapy for patients predicted to be steroid non-responders will further improve outcomes.

*Impact:* The outcome of ASC has substantially improved in the past 25 years, with the potential for further improvement by identifying patients on admission for early escalation of therapy.

## INTRODUCTION

Ulcerative colitis (UC) is a common, chronic, immune-mediated illness, now affecting approximately 1 in 200 of the UK population and increasing in prevalence world-wide.[1,2] Approximately 25% of patients will develop an episode of acute severe colitis[3] (ASC), defined as frequent bloody diarrhoea with evidence of significant systemic biological disturbance (fever, tachycardia, anaemia, elevated inflammatory biomarkers): this complication necessitates hospitalisation for intensive medical management or surgery.[4] Landmark trials in the 20th century demonstrated the efficacy of corticosteroids as first-line medical therapy.[4] The subsequent success of ciclosporin or infliximab as rescue therapy in a proportion of patients with no response to corticosteroids[5-8] led to protocol-driven algorithms of management that have reduced mortality and colectomy rates.[9-14] Nevertheless, consistent data suggest that in the region of 40% of patients with ASC fail to respond to intravenous corticosteroids. [9,10,14]. Predictive models to guide the introduction of medical rescue therapy based on early response to corticosteroids, typically by day three, are in widespread use.[9,10] It remains unclear to what extent this has improved outcomes, or whether earlier identification of steroid non-responders is possible.

We therefore examined outcomes of patients admitted to the Translational Gastroenterology Unit in Oxford, UK, between 2015-9 and compared these with prospective historical data available from the same centre in the pre-biologics’ era (1992-3).[10]

Our further aim was to develop and validate a model on admission to predict which patients were unlikely to respond to steroids, so that rescue therapy or surgery could be expedited. We define and validate in the Oxford population a simple index for risk stratification on the day of admission, involving serum albumin and C-reactive protein (CRP) concentrations, with endoscopic severity, as assessed by the Ulcerative Colitis Endoscopic Index of Severity (UCEIS).[15,16,17] This model, the ADMIT-ASC index, was then successfully validated in independent cohorts in Queensland, Australia and New Delhi, India.

## METHODS

### Study design

A systematic retrospective study of all patients (≥16 years) admitted with ASC in Oxford between 1 May 2015 and 31 October 2019 was performed. Patients were identified from a prospectively maintained endoscopy database (Endobase, Olympus) at Oxford University Hospitals NHS Foundation Trust which comprises two hospitals, John Radcliffe and Horton General Hospital, serving a catchment area population of 700,000. Per local protocol, all patients admitted with suspected ASC underwent flexible sigmoidoscopy on admission, with UCEIS[15,16] recorded in all patients since 2015. This was cross-referenced against the Oxford BRC IBD Biobank database (InfoFlex) to assess completeness of capture. Admission records and corresponding patient case records were then reviewed to confirm patients admitted with ASC during the study period. To ensure consistency and accuracy of data extraction, all patient case records were reviewed by two IBD clinicians (VG and WM).

### Definitions

The <u>diagnosis of UC</u> was confirmed for each patient using established criteria.[18] <u>ASC</u> was defined according to modified Truelove and Witts’ criteria as six or more bloody stools per day with one or more markers of systemic disturbance (heart rate >90, temperature >37.8°C, haemoglobin <105g/L, or CRP >30mg/L).[12] <u>Steroid non-response</u> was defined as administration of medical rescue therapy (ciclosporin or anti-TNF), or colectomy during the same admission. The 1996 <u>Oxford criteria</u> were used to define need for escalation of medical therapy (stool frequency of more than 8 daily, or a stool frequency of 3–8 daily and CRP of more than 45mg/L on the third day after initiating intravenous hydrocortisone).[10]

### Management

Inpatient management followed established Oxford protocol and international guidelines, using intravenous hydrocortisone (100mg four times daily), rectal hydrocortisone (100mg twice daily), intravenous fluids and thromboprophylaxis with low molecular weight heparin.[11,19] Response was assessed on the third day, using the Oxford criteria to determine indications for medical rescue therapy or colectomy.[10] If required, rescue therapy followed hospital protocol (ciclosporin 2mg/kg intravenously until response, then 5mg/kg orally for 3 months; or infliximab 5mg/kg at 0, 2 and 6 weeks, rarely with a higher dose), with the choice based on previous therapy and the judgement of the treating consultant gastroenterologist (Oxford n=3, Gold Coast n=5, Delhi, n=2), if necessary after multidisciplinary discussion. Diagnosis and inpatient management in both validation centres followed the same established guidelines as detailed above, and as in Oxford, in both Australian and Indian centres, protocolised management in ASC was overseen in specialist units, and involved endoscopic assessment on admission, and Day three re-assessment of need for escalation of therapy.[10]

### Outcomes

The primary outcome was steroid non-response during admission with ASC. We separately assessed need for biological therapy, ciclosporin, or colectomy during admission. Re-admission rates with ASC and colectomy rates at 12 months and within the total follow-up period were also assessed. All outcome measures were pre-specified.

### Data collection

In all centres, data were collected on demographic details (gender, age), UC history (disease duration, medication history, extra-intestinal manifestations), admission clinical parameters (stool frequency in preceding 24 hours, heart rate, temperature, haemoglobin, platelet count, white cell count, CRP), admission radiographic findings (presence of toxic megacolon on abdominal radiograph defined by a transverse colon diameter >5.5cm), and endoscopic findings (UCEIS score). Data on use and duration of intravenous steroids, use of medical rescue therapy or need for colectomy during admission were also collected. Following admission, data on re-admission rates with ASC, subsequent requirement for advanced therapy (biologic therapy or tofacitinib) or colectomy, both at 12 months and over the total follow-up period were also collected. Readmissions with ASC were treated as separate events.

### Predictive index development

A predictive index was created using only UK patient data, with only the finally selected model being validated in the Australian and Indian cohorts. Models included a range of up to ten parameters available on admission (age, albumin, CRP, current biologic treatment, disease duration, haemoglobin, platelets, sex, stool frequency, and UCEIS) each over a range of thresholds excluding the top and bottom deciles. One value for CRP and 4 values for albumin were missing. Selection of the final index was based on performance in the UK cohort (ranked by Akaike information criterion), simplicity, and ease of implementation.

### Predictive index validation

For validation of the day one predictive index developed in Oxford, outcomes in two independent cohorts were analysed. Firstly, a systematic retrospective study identified all adult patients coded with ulcerative colitis as the primary indication for admission in electronic patient records and meeting Truelove and Witts’ criteria admitted from 1 January 2015 to 30 April 2020 to Gold Coast University Hospital, Queensland, Australia, serving a catchment area population of approximately 600,000. Secondly, all patients admitted to the All India Institute of Medical Sciences (AIIMS), New Delhi, India, with ASC meeting Truelove and Witts’ criteria between August 2018 and May 2020 (excluding secondary referrals, patients less than 18 years old, and patients who did not receive parenteral steroid treatment) were included. In both these cohorts the index was applied and correlated with outcomes – thereby including 110/128 Australian patients who had a UCEIS score available (all with complete albumin and CRP data), and 62 Indian patients (all with complete albumin, CRP and UCEIS data).

### Statistical analysis

Analysis was performed in R (v3.6). Univariable analysis was performed with Fisher’s exact test for binary variables, and logistic regression for all other variables, and for multivariable logistic regression. P values are given as corrected false discovery rates (FDR).[20] Odds ratios for univariable and multivariable logistic regression are given with confidence intervals calculated using the profile likelihood method [21] and binomial proportion confidence intervals are given for summary statistics of the final model. Preliminary statistical modelling of steroid non-response in the discovery cohort was performed using caret[22] to perform 100 repeats of 10-fold cross-validation.

### Ethical approval

Ethical approval in Oxford was granted through the National Health Service Health Research Authority REC 16/YH/0247 and 09/H1204/30, in Gold Coast by Health Service Human Research Ethics Committee (Ref: LNR/2020/QGC/67173), and in Delhi by AIIMS ethics committee (Ref: IEC-261/04.05.2018)

### Patient and public involvement statement

As this work is a retrospective audit involving examination of routine clinical examinations and clinical outcomes patient and public involvement was not included in the design and conduct of the research.

## RESULTS

### Discovery cohort

In the Discovery cohort, 131 admission events in 117 patients were analysed (42% male, median age 41.2years), including 38 index presentations. No demographic features predicted ASC recurrence (Supplemental Table 1). Demographic and clinical details are given in Tables 1-2.

**Table 1.** Admission demographics in the current Oxford cohort. P values for steroid response vs non-response groups from Wilcoxon rank sum test, or Fisher’s exact test where marked ^F^.

|  | Steroid non- response<br>(n=79) | Steroid response<br>(n=52) |
| --- | --- | --- |
| Gender |  |  |
| Male (n, %) | 34 (43%) | 21 (40%) |
| Female (n, %) | 45 (57%) | 31 (60%) |
| Median age (range), years | 44.6 (16.4-79.6) | 40.6 (16.7-74.3) |
| Age ≥ 60 years (n, %) | 16 (20%) | 9 (17%) |
| Median age at diagnosis (range), years | 29.5 (12.5-79.6) | 30.9 (14.7-73.7) |
| Median disease duration (IQR), years | 1.5 (0.05-7.0) | 0.8 (0.0-7.0) |
| Index presentation as ASC (n, %) | 20 (25%) | 18 (35%) |
| Median follow up (range), weeks | 98.8 (5-213) | 75.4 (10-203) |
| EIM (n, %) | 11 (14%) | 3 (6%) |
| 5-ASA (n, %) |  |  |
| Current | 37 (47%) | 23 (44%) |
| Never | 23 (29%) | 21 (40%) |
| Intolerant/ceased | 19 (24%) | 8 (15%) |
| IM: methotrexate or thiopurine (n, %) |  |  |
| Current |  |  |
| Never | 11 (14%) | 5 (10%) |
| Intolerant/ceased | 49 (62%) | 40 (77%) |
|  | 19 (24%) | 7 (13%) |
| Anti-TNF (n, %) |  |  |
| Current | 3 (4%) | 3 (6%) |
| Never | 67 (85%) | 45 (87%) |
| Intolerant | 4 (5%) | 1 (2%) |
| Primary non-response | 3 (4%) | 1 (2%) |
| Secondary loss of response | 2 (3%) | 1 (2%) |
| Vedolizumab (n, %) |  |  |
| Current | 7 (9%) | 4 (8%) |
| Never | 69 (87%) | 45 (87%) |
| Intolerant | 0 (0%) | 1 (2%) |
| Primary non-response | 1 (1%) | 0 (0%) |
| Secondary loss of response | 2 (3%) | 2 (4%) |
| Tofacitinib |  |  |
| Current | 0 (0%) | 0 (0%) |
| Never | 78 (99%) | 52 (100%) |
| Intolerant | 0 (0%) | 0 (0%) |
| Primary non-response | 1 (1%) | 0 (0%) |
| Secondary loss of response | 0 (0%) | 0 (0%) |
| Previous clinical trial exposure | 1 (1%) | 1 (2%) |
| More than 1 advanced therapy exposure (n, %) | 8 (10%) | 2 (4) |

**5-ASA, 5-aminosalicylate; anti-TNF, anti-tumour necrosis factor; ASC, acute severe colitis; EIM, extra-intestinal manifestations of inflammatory bowel disease; IM, immunomodulator**

**Table 2.** Day 1 admission clinical, laboratory and endoscopic data in 131 admissions for Oxford cohort. P values from Wilcoxon rank sum test, or Fisher’s exact test where marked ^F^.

|  | Steroid non responder (n=79) | Steroid responder (n=52) | p value |
| --- | --- | --- | --- |
| Tachycardia >90 bpm (n, %) | 63 (80%) | 35 (67%) | 0.15<sup>F</sup> |
| Median stool frequency (IQR) | 11 (10-14.5) | 10 (8-13) | 0.19 |
| Median haemoglobin (IQR), g/L | 123 (111.5-134) | 129.5 (108.8-138.3) | 0.33 |
| Median CRP (IQR), mg/L | 101 (55.7-147.7) | 43.4 (23.8-72.9) | 2.7×10<sup>-6</sup> |
| Median albumin (IQR), g/L | 29 (25-32) | 33 (29-35) | 0.33 |
| Median platelet count (IQR), x10<sup>9</sup>/l | 387 (323-468) | 379 (300-441) | 0.50 |
| Toxic megacolon (n, %) | 2 (2.5%) | 0 (0%) | 0.52<sup>F</sup> |
| Truelove and Witts' criteria (n, %) |  |  | 0.006<sup>F</sup> |
| 1 | 18 (23%) | 22 (48%) |  |
| 2 | 40 (51%) | 25 (42%) |  |
| 3 | 20 (25%) | 5 (10%) |  |
| 4 | 1 (1%) | 0 (0%) |  |
| UCEIS score (n, %) |  |  | 0.049<sup>F</sup> |
| 0-2 | 0 (0) | 0 (0) |  |
| 3 | 1 (1%) | 4 (8%) |  |
| 4 | 6 (8%) | 5 (10%) |  |
| 5 | 20 (25%) | 17 (33%) |  |
| 6 | 21 (27%) | 19 (37%) |  |
| 7 | 24 (30%) | 6 (12%) |  |
| 8 | 7 (9%) | 1 (2%) |  |

**bpm, beats per minute; CRP, C-reactive protein UCEIS, Ulcerative Colitis Endoscopic Index of Severity**

Response to corticosteroids without need for medical rescue therapy or surgery was seen in only 52/131 ASC episodes (40%). No demographic parameters were associated with steroid response (Table 1) or ASC recurrence (Supplemental Table 1). Clinical parameters on admission were compared between steroid-responders and non-responders. Median UCEIS was 6 (IQR 5-7) overall, 6 (5-7) for non-responders, and 5.5 (5-6) for responders. Baseline CRP differed between non-responders and responders (median 43mg/L vs 101mg/L, p=2.7 x10^−6^), but albumin, haemoglobin, stool frequency, and platelet count did not (Table 2).

In total, 71 ASC episodes (54%) involved medical rescue therapy, of whom 36 (51%) were given anti-TNF therapy (including 2 who received adalimumab, due to reasons of longer-term intravenous access) and 35 (49%) ciclosporin. At the end of follow-up 60/117 (51%) of patients were receiving advanced therapy (24 infliximab, 9 adalimumab, 21 vedolizumab, 4 tofacitinib, and 2 in clinical trials).

Overall, 19 (15%) patients required colectomy during admission, eight without receiving rescue therapy (2 patient preference, 1 readmission a month following a previous ASC episode, 1 toxic megacolon, and four patients admitted on vedolizumab therapy). Over a median follow-up of 22 (1-49) months, 14 (12%) patients required re-admission with a further episode of ASC.

In 26/131 events (20%) the patient had prior exposure to anti-TNF or vedolizumab (26/93 (28%), excluding index presentations), with 17/131 (13%) events occuring in patients currently on advanced therapy (3 infliximab, 3 adalimumab, 11 vedolizumab). One infliximab-treated patient responded to steroids and continued on infliximab; two received accelerated anti-TNF rescue therapy, neither of whom required surgery. Two of three adalimumab patients responded to intravenous steroids (subsequently discharged on vedolizumab and tofacitinib respectively); the other had surgery without rescue therapy. Of the 11 patients admitted on vedolizumab, 4 responded to intravenous steroids (1 continued vedolizumab, 2 changed to tofacitinib), 3 received rescue therapy (1 ciclosporin, 2 infliximab), and 5 had surgery during the admission (1 after unsuccessful infliximab rescue therapy).

### Comparison of outcomes with pre-biological cohort in Oxford

In comparison to 1992-3[10] (Table 3) the proportion of episodes treated with ciclosporin as rescue therapy was unchanged at 27%, but the proportion of episodes resulting in any medical rescue therapy during admission doubled to 54%, as a result of the use of infliximab in 26% of episodes in the later time period. This coincides with a reduction in colectomy rates during admission from 29% to 15% (p=0.033), and a reduction in readmission with ASC from 35% to 12% (p=0.0017), despite a significantly longer median duration of follow-up, 12 (1992-93) vs 22 months (2015-19, p<0.007).

**Table 3.**
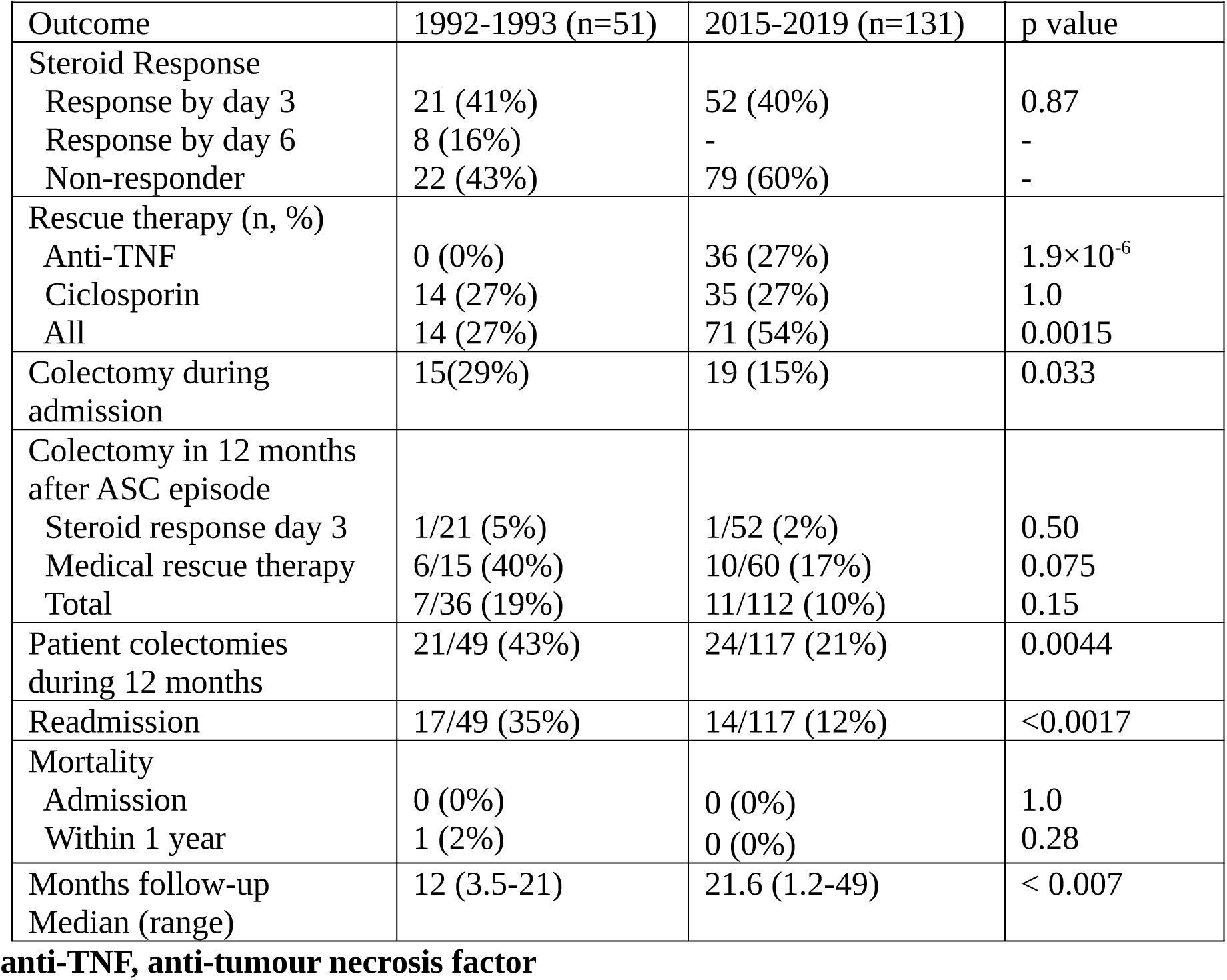
Historical comparison of outcomes in Oxford: P values from Fisher’s exact test, except for follow-up duration which is from a Wilcoxon rank sum test against a worst-case simulated data producing the median and range observed by Travis et al.[10] Colectomy rates for the 1992-3 cohort are from the published follow-up (median 12 months) vs 12 months following discharge in 2015-2019, the difference in follow-up durations is not considered when comparing readmission rates.

In both cohorts the rate of response to intravenous steroids was similar (41% vs 40%, p=0.87). Colectomy rates in the year following admission were approximately eight-fold lower in those who responded to steroid treatment than in those receiving medical rescue therapy (5 vs 40% p=0.013, and 2 vs 17% p=0.010). The overall rate of colectomy for patients within a year fell from 43% to 21% (21/49 in 1993-93 vs 24/117 in 2015-19, p=0.0044).

### Predictive analyses of outcome in the discovery cohort

On univariable analysis, baseline CRP (FDR corrected p=0.00066, OR for a one-unit increase 1.02), albumin (p=0.0066, OR 0.89), UCEIS (p=0.015, OR 1.62), and number of Truelove and Witts’ criteria (p=0.0066, OR 2.43) were significantly associated with steroid non-response (Supplementary Figure 1). Of the individual Truelove and Witts’ criteria, only CRP was independently significant.

Of the components of the UCEIS score, the ‘erosion and ulceration’ component was independently significant (p=0.0066, OR 2.68), and stronger than the overall score. Details of univariable regressions for steroid non-response and rescue therapy are given in Table 4, with further results in Supplemental Table 2.

**Table 4.** Results of univariable analysis in the discovery cohort. Shown as FDR corrected p value (OR, 95% confidence interval), FDR correction was performed for 21 parameters (subcomponents of TW score and UCEIS apart from erosion and ulceration not shown). P values derived from generalized linear models apart from parameters marked with F which were derived from Fisher’s exact test. Steroid non-response denotes rescue therapy or surgery during the admission, rescue therapy includes both anti-TNF treatment and ciclosporin.

|  | Steroid Non-response | Rescue Therapy |
| --- | --- | --- |
| Age | 0.86 (1.00, 0.98-1.02) | 0.26 (1.00, 0.98-1.02) |
| Sex (Male)<sup>F</sup> | 0.86 (1.12, 0.55-2.29) | 0.47 (1.50, 0.75-3.05) |
| Disease Duration | 0.62 (1.01, 0.98-1.05) | 0.81 (0.99, 0.96-1.03) |
| First Admission<sup>F</sup> | 0.70 (0.58, 0.15-1.83) | 0.82 (1.21, 0.39-3.74) |
| Current biologic<sup>F</sup> | 0.82 (1.21, 0.57-2.63) | 0.48 (1.49, 0.71-3.21) |
| Albumin | **0.0066 (0.89, 0.83-0.95)** | **0.029 (0.91, 0.85-0.97)** |
| CRP | **0.00066 (1.02, 1.01-1.03)** | **0.0091 (1.01, 1.01-1.02)** |
| Haemoglobin | 0.60 (0.99, 0.98-1.01) | 0.76 (1.00, 0.98-1.03) |
| Platelets | 0.55 (1.00, 1.00-1.00) | 0.47 (1.00, 1.00-1.01) |
| Stool frequency | 0.53 (1.05, 0.97-1.05) | 0.31 (1.08, 0.99-1.18) |
| TW score | **0.0066 (2.43, 1.45-4.29)** | 0.094 (1.78, 1.10-2.97) |
| TW without CRP | 0.19 (1.75, 0.98-3.23) | 0.46 (1.43, 0.82-2.55) |
| UCEIS score | **0.015 (1.62, 1.18-2.27)** | **0.029 (1.57, 1.15-2.18)** |
| UCEIS erosion & ulceration | **0.0066 (2.68, 1.53-5.00)** | **0.029 (2.13, 1.34-4.17)** |

**CRP, C-reactive protein; TW, Truelove and Witts; UCEIS, Ulcerative Colitis Endoscopic Index of Severity**

In multivariable logistic regression, only CRP was independently significant for steroid non-response (p=0.0076, OR 1.02). Any biologic treatment on admission was associated with the use of rescue therapy with infliximab (p=0.0049, OR 5.65), and not with ciclosporin (p=0.016, OR 0.13). Significant results from multivariable analysis are shown in Supplemental Table 3.

### Predictive models of steroid non-response in the discovery cohort

Steroid non-response was modelled in the discovery cohort with 21 different classification approaches (Supplemental table 4 & supplemental figure 2). Assessment of the importance of each variable in these models revealed CRP to be the most informative variable, followed by UCEIS – in particular the erosion and ulceration component (Supplemental Figure 3). Average prediction accuracy in left out samples from cross-validation ranged from 57% (95% CI 47-66) for K-nearest neighbour (KNN) to 68% (59-77) for multivariate adaptive regression splines (MARS), though across all models accuracy was high for those predicted to be at high or low risk of steroid non-response, and worse for those of intermediate risk. ROC curve for these predictions are shown in Supplemental Figure 4, with mean AUC values ranging from 0.59 for KNN and AdaBoost to 0.75 for MARS, naive Bayes, and sparse partial least squares models. This modelling demonstrates that prediction of steroid response and non-response is possible, but that identifying extremes of risk is more feasible than accurately predicting risk in all patients.

### Selection of a predictive index

Indices were tested in the discovery cohort with a range of thresholds and with both evenly weighted and variably weighted criteria. The final index consists of four points, one each for albumin ≤ 25g/L (19% of patients), and CRP ≥ 100mg/L (34%), with UCEIS ≥ 4 (96%) scoring one point, or two points for UCEIS ≥ 7 (29%). These selected features were all significant in univariable regression (above) and correlations between them are shown in Supplemental Table 5. Of potential fifth components (in decreasing order of utility: stool frequency ≥ 14, haemoglobin ≤ 90g/L, current biologics, platelets ≥500×10^9^/ml, and male sex) only stool frequency made a nominal improvement, but this was not selected for the final model on the basis of the consideration that the added complexity and difficulty of accurately measuring stool frequency outweighed the minor difference in performance (7% improvement in sensitivity at a cost of 4% loss of specificity). The best performing model where UCEIS on admission endoscopy is not available is shown in Supplemental Table 6.

Scores of 3 (14% of patients) and 4 (8%) identified patients with rates of non-response to steroids of 83% and 100% (Table 5). Use of a score threshold ≥3 to identify high risk patients therefore yields a positive predictive value (PPV) of 0.89 (95% CI 0.78-1.00), with an odds ratio (OR) for steroid non-response of 7.6 (6.3-8.8) (Table 6). In comparison positive predictive values for steroid non-response using single-component models of CRP ≥ 100 and albumin ≤ 25 are 0.46 and 0.53 respectively. No significant association exists between current biologic treatment on admission and score (p=0.82), and no correlation was observed between score and non-response to medical rescue therapy.

**Table 5.** Breakdown of steroid response in discovery, and validation cohorts by ADMIT-ASC score involving CRP ≥ 100mg/L (1 point), Albumin ≤ 25g/ L (1 point), UCEIS ≥7 (2 points), or ≥ 4 (1 point).

|  | Discovery (Oxford) |  | Validation (Gold Coast) |  | Validation (India) |  | Combined Validation |  | Total |  |
| --- | --- | --- | --- | --- | --- | --- | --- | --- | --- | --- |
| Score | N | Response % | N | Response % | N | Response % | N | Response % | N | Response % |
| 0 | 3 (2.3%) | 100% | 7 (6.4%) | 100% | 1 (1.6%) | 100% | 8 (4.6%) | 100% | 11 (3.6%) | 100% |
| 1 | 60 (45.8%) | 61.7% | 53 (48.2%) | 73.6% | 35 (56.5%) | 77.1% | 88 (50.1%) | 75.0% | 148 (48.8%) | 70.0% |
| 2 | 40 (30.5%) | 22.5% | 30 (27.3) | 53.3% | 21 (33.9%) | 57.1% | 51 (29.5%) | 54.9% | 91 (30.0%) | 33.0% |
| 3 | 18 (13.7%) | 16.7% | 17 (15.5%) | 17.6% | 5 (8.1%) | 20.0% | 23 (13.3%) | 15.8% | 40 (13.2%) | 17.5% |
| 4 | 10 (7.6%) | 0% | 3 (2.7%) | 0.0% | 0 | 0.0% | 3 (1.7%) | 0.0% | 13 (4.3%) | 0.0% |

**Table 6.** Summary statistics for final model performance predicting steroid non-response using ADMIT-ASC scores of 2-4 as thresholds in the discovery, validation, and combined validation cohorts. Score– threshold score for a prediction of steroid non-response, Prop – proportion of patients at or above threshold, sens – sensitivity, spec – specificity, PPV/NPV – positive/negative predictive value, OR – odds ratio, NNS – number needed to test to prevent one delayed treatment. All statistics show 95% confidence intervals in parentheses. Equivalent results for prediction of steroid response below score 0-2 are shown in Supplemental Table 7, and bootstrapped validation results in Supplemental Table 8.

|  | Discovery (Oxford) |  |  | Validation (Gold Coast) |  |  | Validation (India) |  |  | Combined validation |  |  |
| --- | --- | --- | --- | --- | --- | --- | --- | --- | --- | --- | --- | --- |
| Score | $\geq 2$ | $\geq 3$ | 4 | $\geq 2$ | $\geq 3$ | 4 | $\geq 2$ | $\geq 3$ | 4 | $\geq 2$ | $\geq 3$ | 4 |
| Prop | 51.9% | 21.4% | 7.6% | 45.5% | 18.2% | 2.7% | 41.9% | 8.1% | 0.0% | 44.2% | 14.5% | 1.7% |
| Sens | 0.71<br>(0.61-0.81) | 0.32<br>(0.21-0.42) | 0.13<br>(0.05-0.20) | 0.69<br>(0.55-0.82) | 0.38<br>(0.24-0.52) | 0.07<br>(0.00-0.14) | 0.62<br>(0.41-0.83) | 0.19<br>(0.02-0.36) | - | 0.67<br>(0.55-0.78) | 0.32<br>(0.21-0.43) | 0.05<br>(0.00-0.10) |
| Spec | 0.77<br>(0.65-0.88) | 0.94<br>(0.88-1.00) | 1.00<br>(1.00-1.00) | 0.71<br>(0.60-0.82) | 0.95<br>(0.90-1.00) | 1.00<br>(1.00-1.00) | 0.68<br>(0.54-0.83) | 0.98<br>(0.93-1.00) | - | 0.70<br>(0.61-0.79) | 0.96<br>(0.93-1.00) | 1.00<br>(1.00-1.00) |
| Acc | 0.73<br>(0.66-0.81) | 0.56<br>(0.48-0.65) | 0.47<br>(0.39-0.56) | 0.70<br>(0.61-0.79) | 0.72<br>(0.63-0.80) | 0.62<br>(0.53-0.71) | 0.66<br>(0.41-0.83) | 0.71<br>(0.60-0.82) | - | 0.69<br>(0.62-0.76) | 0.72<br>(0.65-0.78) | 0.63<br>(0.56-0.71) |
| PPV | 0.82<br>(0.73-0.91) | 0.89<br>(0.78-1.00) | 1.00<br>(1.00-1.00) | 0.62<br>(0.49-0.75) | 0.85<br>(0.69-1.00) | 1.00<br>(1.00-1.00) | 0.50<br>(0.31-0.69) | 0.80<br>(0.45-1.00) | - | 0.58<br>(0.47-0.69) | 0.84<br>(0.70-0.98) | 1.00<br>(1.00-1.00) |
| NPV | 0.63<br>(0.52-0.75) | 0.48<br>(0.38-0.57) | 0.43<br>(0.34-0.52) | 0.77<br>(0.66-0.87) | 0.69<br>(0.59-0.78) | 0.61<br>(0.51-0.70) | 0.78<br>(0.64-0.91) | 0.70<br>(0.58-0.82) | - | 0.77<br>(0.69-0.85) | 0.69<br>(0.62-0.77) | 0.63<br>(0.55-0.70) |
| OR | 8.1<br>(7.3-8.9) | 7.6<br>(6.3-8.8) | Inf | 5.4<br>(4.5-6.2) | 12.5<br>(11.2-13.9) | Inf | 3.5<br>(2.4-4.6) | 9.4<br>(7.1-11.7) | - | 4.6<br>(4.0-5.3) | 11.9<br>(10.8-13.0) | Inf |
| NNS | 2.3<br>(2.0-2.9) | 5.2<br>(3.9-8.1) | 13.1<br>(8.2-32.4) | 3.5<br>(2.7-5.1) | 6.5<br>(4.5-11.5) | 36.7<br>(17.3-315.9) | 4.8<br>(3.2-9.2) | 15.5<br>(8.0-297.3) | - | 3.9<br>(3.1-5.2) | 8.2<br>(5.8-13.7) | 57.3<br>(27.0-471.1) |
| AUC | 0.74<br>(0.65-0.83) | 0.63<br>(0.53-0.73) | 0.56<br>(0.46-0.66) | 0.70<br>(0.60-0.80) | 0.67<br>(0.57-0.77) | 0.53<br>(0.42-0.64) | 0.65<br>(0.51-0.79) | 0.58<br>(0.44-0.73) | - | 0.68<br>(0.60-0.76) | 0.64<br>(0.56-0.72) | 0.52<br>(0.43-0.61) |

### Validation Cohorts

At the Gold Coast University Hospital, a total of 128 patients (50% male, median age 35 years) presented with ASC between January 1^st^, 2015 and April 30^th^, 2020. Demographics and clinical parameters are given in Supplemental Tables <u>9 & 10</u>. At presentation, 12 (9%) patients were on biological therapy (8 anti-TNF, 4 vedolizumab), and for 41 patients (32%) the admission was their index presentation of UC.

Medical rescue therapy was given to 51 (40%) patients (4 ciclosporin, 47 infliximab). Ten (8%) patients had a colectomy during admission, with one direct colectomy. As in the discovery cohort, all features selected for the final index were significant in univariable regression (Supplemental Table 11). Validation of the index was performed in the 110 (86%) patients with a UCEIS score recorded (all with CRP and albumin results). There were no significant differences between this subset and the complete Gold Coast cohort (Supplemental Table 1).

At the All India Institute of Medical Science, 62 patients (40% male, median age 35.5 years) presented with ASC between August 2018 and May 2020. Demographics and clinical parameters are given in supplemental tables 12 & 13. Medical rescue therapy was given to 17 (27%) patients (2 ciclosporin, 15 anti-TNF) and 7 (11%) patients had a colectomy during admission.

When the final model was applied to the Australian cohort with available UCEIS scores (n=110), a similar proportion of patients scored ≥3 (18% vs 21%, p=0.63); in the Indian cohort this was significantly lower (8%, p=4.6×10^−7^). However steroid response rates for each score were similar across all three cohorts (Table 5), apart from patients with the intermediate scores of 1 or 2 who were more likely to receive rescue therapy or surgery in the UK cohort (p=0.10 & 0.002 respectively). A score of 0 was100% predictive of steroid response and a score of 4 100% predictive of steroid non-response across all three cohorts.

### Proposed implementation from pooled results

Although a score of 4 was 100% predictive of steroid non-response, only 13 patients (4.3%) scored 4. Extending the threshold to a score of 3 increased the proportion of patients to 17.5%. In the combined validation cohorts this threshold is highly specific (0.96, 95% CI 0.93-1.00), with a PPV of 0.84 (0.70-0.98) and OR of 11.9 (10.8-13.0). Use of this threshold to advance treatment would prevent one delayed treatment for every 8.2 (5.8-13.7) patients assessed. Summary statistics in the discovery cohort and both validation cohorts separately and combined are shown in Table 6.

## DISCUSSION

This study provides new insights into the management and outcome of severe ulcerative colitis with relevance to current clinical practice. We demonstrate that urgent colectomy rates have halved from 29% to 15% in 2015-19, in comparison with the 1992-3 series from Oxford.[10] Re-admission and colectomy rates after discharge have also fallen by two-thirds and a half respectively. The risks of colectomy in those without a clear steroid response remain 8-fold elevated in the year following discharge.[10,23] The early identification of this group with initiation and maintenance of longer-term effective treatment therefore remains a priority beyond management of the episode of ASC.

The study also identifies a predictive index for steroid non-response that can be applied on the day of admission, validated in two independent cohorts from separate continents. A combination of CRP ≥ 100mg/L, albumin ≤ 25g/L, and a UCEIS score of 7-8 (2 points) is associated with a Day 1 score of 4 and certain likelihood of steroid non-response (100% in both discovery and two validation cohorts), regardless of whether the patient is already established on biologics. A score of 3-4 is associated with an 84% chance of non-response to steroids and we propose a score of 3 or greater in a biological-naïve patient is an indication for early medical rescue therapy. In a patient admitted on biological therapy a score of 3 mandates surgical discussion early in admission. The case for early medical rescue therapy is clear, but the question of whether steroid treatment should continue concomitantly needs future study.

The historical perspective from the same centre is informative. A key difference between the 1992-93 study from Oxford and the present study is the precise doubling of the number receiving rescue medical therapy. In both datasets, an equal proportion of patients received ciclosporin (27%). In the current Gold Coast dataset, where approximately 40% of patients received rescue therapy with ciclosporin or infliximab, we report an acute colectomy rate of <10%. In the Indian cohort, the colectomy rate was 11%, with 27% given rescue therapy. This provides compelling evidence that alterations in treatment strategy in the modern era, including - but not exclusively - intervention with biological therapies, have improved clinical outcome in ASC, compared with historical outcomes. Standardised protocols for assessing response, guiding care and emphasis on multi-disciplinary team involvement appear fundamentally important to set standards for care, inform patient discussion and as a basis for further audit.

The current study confirms previous observations from ourselves and others that the biological severity on admission, as defined by Truelove and Witts, predicts resistance to therapy[23-28]. Our data suggest that CRP is the key parameter associated with predicting non-response to steroids.

By careful modelling a simple clinical index, ADMIT-ASC, assessed on admission was derived from the CRP, albumin and a validated index of endoscopic severity (UCEIS). The index reliably identifies a subset of approximately 1 in 5 patients with highly active disease, who will fail to show response to parenteral corticosteroid therapy[11] and require consideration of escalation of therapy or surgery. These data provide a rationale to refine the paradigm for management, which is currently predicated on treating all patients with parenteral steroids and assessing steroid response on day 3.

As a counterpoint, those patients with a score of 2 or less may be managed by conventional approach, with reassessment of steroid response, and need for escalation at day 3. A score of 0 may be particularly reassuring, since we found complete concordance with steroid response across the cohorts.

Defining risk on the day of admission carries advantages in decision-making, contingency planning, and patient counselling, especially in those at high-risk of steroid non-response. The ADMIT-ASC index is reproducible and defines high-risk patients accurately. The index is simple and implementable without other changes in practice. All the parameters are readily and reproducibly defined, without the awkward need for assessment of stool frequency. Our data break new ground by involving contemporary discovery and validation cohorts who have been managed using biologics in induction or maintenance and by taking advantage of a validated endoscopic scoring system, notwithstanding geographical, and ethnic differences between all three populations.

There are, of course, limitations. The study is retrospective in design and did not allow reliable assessment of pre-admission therapy in the discovery cohort. Nonetheless, patient identification appeared complete and painstaking searches of available databases and paper case notes were performed to avoid ascertainment bias. The validity of the discovery dataset was confirmed by independent replication in two datasets from different continents. This is the first time a predictive index in ASC has been so validated.

We did not formally capture disease morbidity or complications of treatment, other than in the context of colectomy rates or re-admission. These are subject to regular audit and there is no signal of complication rates associated either with drug therapy or colectomy.[29] We also recognise that although the proposed index accurately identifies patients at a very high or low likelihood of response to steroid treatment, there remains a significant proportion who score 1 or 2 and in whom steroid responsiveness is difficult to predict. The study was performed in three specialist centres, where management was the responsibility of IBD-focused clinicians, who followed protocols to optimise outcomes developed in Oxford and adopted internationally. Nevertheless, most (80%) of patients came from the local population, as in any secondary care hospital. The replication in two cohorts from different genetic backgrounds, previous treatment, environmental exposures, age and disease duration is a particular strength of our study.

The present findings are highly relevant to daily practice by clinicians involved in the care of patients with acute severe colitis. The validation of an index that relies on two routine blood tests and assessment of endoscopic severity on admission is a guide to early stage decision-making. We believe that this predictive model can appropriately be applied to trials and clinical practice.

## Data Availability

Please contact the corresponding authors for access to anonymised data.

## Acknowledgements

The authors acknowledge the clinical and academic contributions made to this work by Professor Satish Keshav, who sadly passed away before the study was completed.

This work is funded by the Oxford NIHR Biomedical Research Centre - Gastroenterology and Mucosal Immunology Theme. The views expressed are those of the authors and not necessarily those of the NHS, the NIHR or the Department of Health.

We acknowledge the contribution of the Oxford IBD cohort study BRC, which is supported by the NIHR Oxford Biomedical Research Centre, University of Oxford. The authors thank Sister and staff of the specialist wards, IBD Specialist Nurses and administrative staff, as well as our patients. The authors are also grateful to the other Oxford IBD Cohort Investigators: Carolina V. Arancibia-Cárcamo, Adam Bailey, Ellie Barnes, Elizabeth Bird-Lieberman, Barbara Braden, Jane Collier, James East, Alessandra Geremia, Lucy Howarth, Satish Keshav, Paul Klenerman, Simon Leedham, Fiona Powrie, Astor Rodrigues, Peter Sullivan and Holm Uhlig.

## Data sharing statement

Please contact the corresponding authors for access to anonymised data.

## Author contributions

JS proposed the study. WM, VG, DS, PRK, SKV, and SK collected the clinical data. AA and PKR performed the analysis. AA, JS, VG, WM & TC contributed to study design and prepared the first draft, and all authors interpreted the results, and reviewed and contributed to the final manuscript.

## REFERENCES

1 Ungaro R, Mehandru S, Allen PB, et al. Ulcerative colitis. Lancet 2017;389:1756–70. doi:10.1016/S0140-6736(16)32126-2

2 Ng SC, Shi HY, Hamidi N, et al. Worldwide incidence and prevalence of inflammatory bowel disease in the 21st century: a systematic review of population-based studies. Lancet 2017;390:2769–78. doi:10.1016/S0140-6736(17)32448-0

3 Travis S, Satsangi J, Lémann M. Predicting the need for colectomy in severe ulcerative colitis: A critical appraisal of clinical parameters and currently available biomarkers. Gut 2011;60:3–9. doi:10.1136/gut.2010.216895

4 Truelove SC, Witts LJ. Cortisone in Ulcerative Colitis. BMJ 1955;2:1041–8. doi:10.1136/bmj.2.4947.1041

5 Lichtiger S, Present DH, Kornbluth A, et al. Cyclosporine in Severe Ulcerative Colitis Refractory to Steroid Therapy. N Engl J Med 1994;330:1841–5. doi:10.1056/NEJM199406303302601

6 Järnerot G, Hertervig E, Friis-Liby I, et al. Infliximab as rescue therapy in severe to moderately severe ulcerative colitis: A randomized, placebo-controlled study. Gastroenterology 2005;128:1805–11. doi:10.1053/j.gastro.2005.03.003

7 Williams JG, Alam MF, Alrubaiy L, et al. Infliximab versus ciclosporin for steroid-resistant acute severe ulcerative colitis (CONSTRUCT): a mixed methods, open-label, pragmatic randomised trial. Lancet Gastroenterol Hepatol 2016; 1: 15–24. doi:10.1016/S2468-1253(16)30003-6

8 Laharie D, Bourreille A, Branche J, et al. Ciclosporin versus infliximab in patients with severe ulcerative colitis refractory to intravenous steroids: A parallel, open-label randomised controlled trial. Lancet 2012;380:1909–15. doi:10.1016/S0140-6736(12)61084-8

9 Ho GT, Mowat C, Goddard CJR, et al. Predicting the outcome of severe ulcerative colitis: Development of a novel risk score to aid early selection of patients for second-line medical therapy or surgery. Aliment Pharmacol Ther 2004; 19: 1079–87. doi:10.1111/j.1365-2036.2004.01945.x

10 Travis SPL, Farrant JM, Ricketts C, et al. Predicting outcome in severe ulcerative colitis. Gut 1996;38:905–10. doi:10.1136/gut.38.6.905

11 Lamb CA, Kennedy NA, Raine T, et al. British Society of Gastroenterology consensus guidelines on the management of inflammatory bowel disease in adults. Gut 2019;68:s1–106. doi:10.1136/gutjnl-2019-318484

12 Harbord M, Eliakim R, Bettenworth D, et al. Third European evidence-based consensus on diagnosis and management of ulcerative colitis. Part 2: Current management. J Crohn’s Colitis 2017;11:769–84. doi:10.1093/ecco-jcc/jjx009

13 Singh S, Allegretti JR, Siddique SM, Terdiman JP. AGA Technical Review on the Management of Moderate to Severe Ulcerative Colitis. Gastroenterology 2020;158:1465–1496. doi:10.1053/j.gastro.2020.01.007

14 Grant RK, Jones G-R, Plevris N, et al. The ACE (Albumin, CRP and Endoscopy) Index in Acute Colitis: A Simple Clinical Index on Admission that Predicts Outcome in Patients With Acute Ulcerative Colitis. Inflamm Bowel Dis 2021;27:451–7. doi:10.1093/ibd/izaa088

15 Travis SPL, Schnell D, Krzeski P, et al. Developing an instrument to assess the endoscopic severity of ulcerative colitis: The Ulcerative Colitis Endoscopic Index of Severity (UCEIS). Gut 2012;61:535–42. doi:10.1136/gutjnl-2011-300486

16 Travis SPL, Schnell D, Krzeski P, et al. Reliability and initial validation of the ulcerative colitis endoscopic index of severity. Gastroenterology 2013;145:987–95. doi:10.1053/j.gastro.2013.07.024

17 Gupta V, Mohsen W, Chapman TP, Satsangi J. Predicting Outcome in Acute Severe Colitis— Controversies in Clinical Practice in 2021. J Crohn’s Colitis 2021;15:1211–21. doi:10.1093/ecco-jcc/jjaa265

18 Lennard-Jones JE. Classification of Inflammatory Bowel Disease. Scand J Gastroenterol 1989;24:2–6. doi:10.3109/00365528909091339

19 Jakobovits SL, Travis SPL. Management of acute severe colitis. Br Med Bull 2005;75–76:131– 44. doi:10.1093/bmb/ldl001

20 Benjamini Y, Hochberg Y. Controlling the false discovery rate: a practical and powerful approach to multiple testing. J R Stat Soc Ser B 1995;57:289–300. https://www.jstor.org/stable/2346101

21 Venzon DJ, Moolgavkar SH. A Method for Computing Profile-Likelihood-Based Confidence Intervals. Appl Stat 1988;37:87. doi:10.2307/2347496

22 Kuhn M. Building predictive models in R using the caret package. J Stat Softw 2008;28:1–26. doi:10.18637/jss.v028.i05

23 Bojic D, Radojicic Z, Nedeljkovic-Protic M, et al. Long-term outcome after admission for acute severe ulcerative colitis in Oxford: The 1992-1993 cohort. Inflamm Bowel Dis 2009;15:823–8. doi:10.1002/ibd.20843

24 Dinesen LC, Walsh AJ, Protic MN, et al. The pattern and outcome of acute severe colitis. J Crohn’s Colitis 2010;4:431–7. doi:10.1016/j.crohns.2010.02.001

25 Lynch RW, Lowe D, Protheroe A, et al. Outcomes of rescue therapy in acute severe ulcerative colitis: data from the United Kingdom inflammatory bowel disease audit. Aliment Pharmacol Ther 2013;38:935–45. doi:10.1111/apt.12473

26 Henriksen M, Jahnsen J, Lygren I, et al. C-reactive protein: a predictive factor and marker of inflammation in inflammatory bowel disease. Results from a prospective population-based study. Gut 2008;57:1518–23. doi:10.1136/gut.2007.146357

27 Stallmach A, Nickel A, Lehmann T, et al. Parameters of a severe disease course in ulcerative colitis. World J Gastroenterol 2014;20:12754–80. doi:10.3748/wjg.v20.i35.12574

28 Jain S, Kedia S, Bopanna S, et al. Are Truelove and Witts criteria for diagnosing acute severe colitis relevant for the Indian population? A prospective study. Intest Res 2018;16:69–74. doi:10.5217/ir.2018.16.1.69

29 Hyde GM, Jewell DP, Kettlewell MGW, Mc. C. Mortensen NJ. Cyclosporin for severe ulcerative colitis does not increase the rate of perioperative complications. Dis Colon Rectum 2001;44:1436–40. doi:10.1007/BF02234594

