## Supplemental tables and figures for "Optimising Early Management of Acute Severe Ulcerative Colitis in the Biologics Era: Admission Model for Intensification of Therapy in Acute Severe Colitis (ADMIT–ASC)"

### Supplemental Data

### **Supplemental Table 1** *Admission demographics*

| | Oxford | | Gold Coast | |
|---|---|---|---|---|
| | Patients (at first admission) (n=117) | Total Admissions (n=131) | Model replication cohort (n=110) | Total admissions (n=128) |
| Gender<br>Male (n, %)<br>Female (n, %) | <br>49 (42%)<br>68 (58%) | <br>55 (42%)<br>76 (58%) | <br>54 (49%)<br>56 (51%) | <br>64 (50%)<br>64 (50%) |
| Median age (range), years | 41.2 (16.4-79.6) | 41.2 (16.4-79.6) | 34.5 (18-88) | 35 (18-88) |
| Age ≥ 60 years (n, %) | 22 (19%) | 25 (19%) | 22 (20%) | 26 (20%) |
| Median age at diagnosis (range), years | 31.2 (12.5-79.6) | 30.6 (12.5-79.6) | 28.5 (10-80) | 29 (10-80) |
| Median disease duration (IQR), years | 1.0 (0.0-6.0) | 1.0 (0.0-7.0) | 1.75 (0-6) | 2 (0-6) |
| Index presentation as ASC (n, %) | 38 (32%) | 38 (29%) | 35 (32%) | 41 (32%) |
| Median follow up post ASC (range), weeks | 98.8 (5-213) | 93.6 (5-213) | 78.5 (7-280) | 76.5 (7-280) |
| EIM (n, %) | 12 (10%) | 14 (11%) | 13 (12%) | 14 (11%) |
| 5-ASA (n, %)<br>Current<br>Never<br>Intolerant/ceased | <br>56 (48%)<br>42 (36%)<br>19 (16%) | <br>60 (46%)<br>44 (34%)<br>27 (21%) | <br>53 (48%)<br>35 (32%)<br>22 (20%) | <br>65 (51%)<br>39 (30%)<br>24 (19%) |
| IM: methotrexate or thiopurine (n, %)<br>Current<br>Never<br>Intolerant/ceased | <br>12 (10%)<br>82 (70%)<br>23 (20%) | <br>16 (12%)<br>89 (68%)<br>26 (20%) | <br>12 (11%)<br>80(73%)<br>18 (16%) | <br>14 (11%)<br>86 (67%)<br>28 (22%) |
| Anti-TNF (n, %)<br>Current<br>Never<br>Intolerant<br>Primary non-response<br>Secondary loss of response | <br>4 (3%)<br>110 (94%)<br>2 (2%)<br>4 (3%)<br>3 (3%) | <br>6 (5%)<br>122 (93%)<br>3 (2%)<br>6 (5%)<br>4 (3%) | <br>8 (7%)<br>95 (86%)<br>4 (4%)<br>0<br>3 (3%) | <br>8 (6%)<br>110 (86%)<br>5 (4%)<br>0<br>5 (4%) |
| Vedolizumab (n, %)<br>Current<br>Never<br>Intolerant<br>Primary non-response<br>Secondary loss of response | <br>5 (4%)<br>108 (92%)<br>1 (1%)<br>0 (0%)<br>3 (3%) | <br>11 (8%)<br>114 (87%)<br>1 (1%)<br>1 (1%)<br>4 (3%) | <br>4 (4%)<br>105 (95%)<br>0<br>0<br>1 (1%) | <br>4 (3%)<br>122 (95%)<br>0<br>0<br>2 (2%) |
| Tofacitinib<br>Current<br>Never<br>Intolerant<br>Primary non-response<br>Secondary loss of response | <br>0 (0%)<br>117 (100%)<br>0 (0%)<br>0 (0%)<br>0 (0%) | <br>0 (0%)<br>130 (99%)<br>0 (0%)<br>1 (1%)<br>0 (0%) | 0 | 0 |
| Previous clinical trial exposure | 2 (2%) | 2 (2%) | 0 | 0 |
| More than 1 advanced therapy exposure (n, %) | 7 (6%) | 10 (8%) | 3 (3%) | 4 (3%) |

**Supplemental Table 2** *Univariable logistic regression results for other outcomes in the discovery cohort. Shown as FDR corrected p value (OR, 95% confidence interval), FDR correction was performed for 21 parameters (subcomponents of TW score and UCEIS apart from erosion and ulceration not shown). P values derived from generalized linear models apart from parameters marked with * which were derived from Fisher's exact test.*

| | Colectomy during admission | Colectomy within 1 year | Rescue Therapy (anti-TNF) | Rescue Therapy (Ciclosporin) |
|---|---|---|---|---|
| Age | 0.78 (1.01, 0.98-1.04) | 0.73 (1.01, 0.99-1.03) | 0.26 (0.98, 0.96-1.00) | 0.45 (1.02, 1.00-1.04) |
| Sex (Male)* | 0.76 (0.59, 0.20-1.62) | 0.31 (0.51, 0.21-1.20) | 0.73 (1.34, 0.62-2.91) | 0.73 (.123, 0.56-2.69) |
| Disease Duration | 0.61 (1.03, 0.98-1.07) | 0.19 (1.04, 1.00-1.08) | 0.40 (0.97, 0.92-1.01) | 0.55 (1.02, 0.98-1.06) |
| First Admission* | 0.19 (0.24, 0.07-0.89) | 0.26 (0.34, 0.11-1.13) | 0.40 (0.46, 0.15-1.50) | 0.47 (5.33, 1.00-98.62) |
| Current biologic* | 0.14 (0.10, 0.01-0.51) | **0.0016 (0.05, 0.0-0.26)** | **0.00029 (6.29, 2.76-14.88)** | **0.011 (0.14, 0.03-0.44)** |
| Albumin | 0.36 (0.93, 0.86-1.01) | 0.082 (0.90, 0.84-0.97) | 0.67 (0.97, 0.91-1.04) | 0.081 (0.91, 0.85-0.98) |
| CRP | 0.19 (1.01, 1.00-1.01) | 0.19 (1.01, 1.00-1.01) | 0.38 (1.00, 1.00-1.01) | 0.081 (1.01, 1.00-1.01) |
| Haemoglobin | 0.86 (1.00, 0.98-1.03) | 0.73 (1.00, 0.98-1.02) | 0.26 (0.98, 0.96-1.00) | 0.54 (1.01, 0.99-1.04) |
| Platelets | 0.68 (1.00, 1.00-1.01) | 0.73 (1.00, 1.00-1.01) | 0.90 (1.00, 1.00-1.00) | 0.55 (1.00, 1.00-1.01) |
| Stool frequency | 0.76 (1.04, 0.93-1.15) | 0.97 (1.00, 0.90-1.09) | 0.71 (1.03, 0.94-1.12) | 0.54 (1.05, 0.96-1.15) |
| TW score | 0.19 (2.06, 1.06-4.15) | 0.19 (1.86, 1.06-3.34) | 0.26 (1.7, 1.00-2.94) | 0.67 (1.18, 0.69-2.00) |
| TW without CRP | 0.21 (2.23, 1.01-5.13) | 0.20 (1.94, 1.00-1.39) | 0.40 (1.51, 0.81-2.88) | 0.93 (1.03, 0.55-1.92( |
| UCEIS score | 0.86 (0.92, 0.61-1.39) | 0.73 (1.11, 0.79-1.59) | 0.26 (1.4, 1.00-2.01) | 0.54 (1.24, 0.89-1.76) |
| UCEIS erosion & ulceration | 0.93 (1.08, 0.52-2.27) | 0.31 (1.59, 0.86-3.07) | **0.039 (2.61, 1.40-5.13)** | 0.73 (1.12, 0.63-2.02) |

**Supplemental Table 3** *Significant results from multivariable analysis in the discovery cohort (P values uncorrected)*

| Outcome | Parameter | P | OR (95% CI) |
|---|---|---|---|
| Rescue Therapy (Ciclosporin) | Current Biologic | 0.016 | 0.13 (0.02-0.58) |
| Rescue Therapy (Anti-TNF) | Hb | 0.0025 | 0.92 (0.87-0.97) |
| Rescue Therapy (Anti-TNF) | Current Biologic | 0.0049 | 5.65 (1.75-20.05) |
| Colectomy during admission | First Admission | 0.032 | 0.1 (0.01-0.77) |
| Colectomy during admission | Current Biologic | 0.038 | 0.06 (0-0.55) |
| Colectomy Within 1 year | Current Biologic | 0.0022 | 0.02 (0-0.15) |
| Steroid Non-response | CRP | 0.0076 | 1.02 (1.01-1.03) |

**Supplemental Table 4:** *Summary statistics (95% CI): PPV – positive predictive value, NPV – negative predictive value, OR – odds ratio, PLR – positive likelihood ratio, NLR – negative likelihood ratio, $F_{0.5}$ – F-score with β = 0.5. Models: BLR - Boosted Logistic Regression, LDA - Linear Discriminant Analysis, MARS - Multivariate Adaptive Regression Spline, BCT - Boosted Classification Trees, CART – Classification And Regression Trees, NSC - Nearest Shrunken Centroids, PLS - Partial least squares, QDA - Quadratic Discriminant Analysis, KNN - K-Nearest Neighbours*

| model | Sensitivity | Specificity | Accuracy | PPV | NPV | OR | PLR | NLR | $F_{0.5}$ |
|---|---|---|---|---|---|---|---|---|---|
| BLR | 0.50 (0.35-0.66) | 0.67 (0.55-0.79) | 0.60 (0.51-0.70) | 0.50 (0.34-0.66) | 0.67 (0.55-0.79) | 2.0 (1.2-2.9) | 1.5 (0.6-2.5) | 0.7 (0.2-1.3) | 0.50 (0.40-0.60) |
| Random Forrest | 0.54 (0.39-0.70) | 0.69 (0.57-0.80) | 0.63 (0.54-0.73) | 0.53 (0.38-0.69) | 0.70 (0.58-0.81) | 2.6 (1.8-3.5) | 1.7 (0.7-2.8) | 0.7 (0.2-1.1) | 0.54 (0.44-0.63) |
| Sparse LDA | 0.07 (0.00-0.15) | 0.96 (0.91-1.00) | 0.61 (0.51-0.70) | 0.52 (0.10-0.95) | 0.61 (0.51-0.71) | 1.7 (0.0-3.5) | 1.7 (1.3-2.0) | 1.0 (0.1-1.9) | 0.23 (0.14-0.31) |
| MARS | 0.65 (0.50-0.80) | 0.70 (0.58-0.81) | 0.68 (0.59-0.77) | 0.59 (0.44-0.73) | 0.75 (0.64-0.86) | 4.3 (3.4-5.1) | 2.1 (0.8-3.5) | 0.5 (0.2-0.8) | 0.60 (0.50-0.69) |
| BCT | 0.58 (0.43-0.74) | 0.68 (0.56-0.80) | 0.64 (0.55-0.73) | 0.54 (0.39-0.69) | 0.71 (0.59-0.83) | 2.9 (2.1-3.8) | 1.8 (0.7-3.0) | 0.6 (0.2-1.0) | 0.55 (0.45-0.65) |
| AdaBoost | 0.49 (0.33-0.65) | 0.77 (0.66-0.88) | 0.66 (0.57-0.75) | 0.58 (0.42-0.75) | 0.70 (0.59-0.81) | 3.2 (2.4-4.1) | 2.1 (0.9-3.4) | 0.7 (0.2-1.1) | 0.56 (0.47-0.66) |
| CART | 0.61 (0.46-0.76) | 0.65 (0.53-0.77) | 0.63 (0.54-0.73) | 0.53 (0.39-0.68) | 0.72 (0.60-0.84) | 2.9 (2.1-3.7) | 1.7 (0.6-2.9) | 0.6 (0.2-1.0) | 0.55 (0.45-0.65) |
| LDA | 0.50 (0.35-0.66) | 0.68 (0.57-0.80) | 0.61 (0.52-0.71) | 0.51 (0.36-0.67) | 0.68 (0.56-0.79) | 2.2 (1.4-3.0) | 1.6 (0.6-2.6) | 0.7 (0.2-1.2) | 0.51 (0.41-0.61) |
| LDA (2) | 0.52 (0.37-0.68) | 0.68 (0.56-0.80) | 0.62 (0.52-0.71) | 0.52 (0.36-0.67) | 0.68 (0.57-0.80) | 2.3 (1.5-3.2) | 1.6 (0.6-2.7) | 0.7 (0.2-1.2) | 0.52 (0.42-0.62) |
| Stepwise LDA | 0.54 (0.38-0.69) | 0.76 (0.65-0.87) | 0.67 (0.58-0.76) | 0.59 (0.43-0.75) | 0.71 (0.60-0.82) | 3.6 (2.8-4.5) | 2.2 (0.9-3.5) | 0.6 (0.2-1.0) | 0.58 (0.48-0.68) |
| Naive Bayes | 0.58 (0.43-0.74) | 0.72 (0.60-0.83) | 0.66 (0.57-0.76) | 0.57 (0.42-0.73) | 0.72 (0.61-0.84) | 3.5 (2.7-4.3) | 2.0 (0.8-3.3) | 0.6 (0.2-1.0) | 0.58 (0.48-0.67) |
| Naive Bayes (2) | 0.48 (0.33-0.64) | 0.78 (0.67-0.88) | 0.66 (0.57-0.75) | 0.59 (0.42-0.76) | 0.70 (0.59-0.81) | 3.3 (2.4-4.2) | 2.2 (1.0-3.4) | 0.7 (0.2-1.1) | 0.57 (0.47-0.66) |
| NSC | 0.55 (0.40-0.71) | 0.75 (0.64-0.86) | 0.67 (0.58-0.77) | 0.60 (0.44-0.75) | 0.72 (0.61-0.83) | 3.8 (2.9-4.6) | 2.2 (0.9-3.6) | 0.6 (0.2-1.0) | 0.59 (0.49-0.68) |
| C5.0 | 0.61 (0.46-0.76) | 0.65 (0.53-0.77) | 0.64 (0.54-0.73) | 0.54 (0.39-0.68) | 0.72 (0.60-0.84) | 2.9 (2.1-3.7) | 1.8 (0.6-2.9) | 0.6 (0.2-1.0) | 0.55 (0.45-0.65) |
| DeepBoost | 0.57 (0.42-0.72) | 0.69 (0.57-0.81) | 0.64 (0.55-0.74) | 0.55 (0.39-0.70) | 0.71 (0.59-0.82) | 2.9 (2.1-3.8) | 1.8 (0.7-3.0) | 0.6 (0.2-1.0) | 0.55 (0.45-0.65) |
| PLS | 0.55 (0.40-0.71) | 0.75 (0.64-0.86) | 0.67 (0.58-0.76) | 0.59 (0.43-0.75) | 0.72 (0.61-0.83) | 3.7 (2.9-4.6) | 2.2 (0.9-3.5) | 0.6 (0.2-1.0) | 0.58 (0.49-0.68) |
| Sparse PLS | 0.54 (0.38-0.69) | 0.76 (0.65-0.87) | 0.67 (0.58-0.76) | 0.60 (0.44-0.76) | 0.71 (0.60-0.82) | 3.7 (2.8-4.5) | 2.2 (0.9-3.6) | 0.6 (0.2-1.0) | 0.58 (0.49-0.68) |
| QDA | 0.41 (0.26-0.56) | 0.72 (0.60-0.83) | 0.59 (0.50-0.69) | 0.49 (0.32-0.66) | 0.65 (0.53-0.76) | 1.7 (0.9-2.6) | 1.4 (0.6-2.3) | 0.8 (0.2-1.4) | 0.47 (0.37-0.57) |
| KNN | 0.54 (0.39-0.70) | 0.58 (0.46-0.71) | 0.57 (0.47-0.66) | 0.46 (0.32-0.60) | 0.66 (0.53-0.79) | 1.7 (0.8-2.5) | 1.3 (0.4-2.2) | 0.8 (0.3-1.3) | 0.48 (0.38-0.57) |
| Neural Network | 0.57 (0.42-0.73) | 0.67 (0.55-0.79) | 0.63 (0.54-0.73) | 0.53 (0.38-0.68) | 0.70 (0.59-0.82) | 2.7 (1.9-3.5) | 1.7 (0.6-2.8) | 0.6 (0.2-1.1) | 0.54 (0.44-0.64) |
| Elastic Net | 0.50 (0.35-0.66) | 0.78 (0.68-0.89) | 0.67 (0.58-0.76) | 0.60 (0.43-0.77) | 0.70 (0.59-0.81) | 3.6 (2.7-4.5) | 2.3 (1.0-3.6) | 0.6 (0.2-1.1) | 0.58 (0.48-0.68) |

**Supplemental Table 5** *Spearman correlations between ADMIT-ASC components in the discovery cohort*

| | | ρ | P value |
|---|---|---|---|
| CRP | Albumin | -0.44 | 2.3×10-7 |
| CRP | UCEIS | 0.24 | 0.006 |
| Albumin | UCEIS | -0.19 | 0.03 |

**Supplemental Table 6** The best performing model in the discovery cohort if UCEIS is not available consists of 1 point for each of CRP ≥ 100, albumin ≤ 25, and haemoglobin ≤ 120. A) *Summary statistics for model performance predicting steroid non-response using scores of 0-3 as thresholds in the validation cohort. PPV/NPV – positive/negative predictive value, OR – odds ratio. All statistics show 95% confidence intervals in parentheses. B) Number and steroid non-response rate for each score group.*

A

| Threshold | 0 | ≥ 1 | ≥ 2 | 3 |
|---|---|---|---|---|
| Proportion of patients | 45 (40.9%) | 65 (59.1%) | 24 ( 21.8%) | 5 (4.5%) |
| Sensitivity | 0.24 (0.13-0.39) | 0.75 (0.60-0.87) | 0.33 (0.20-0.49) | 0.09 (0.02-0.21) |
| Specificity | 0.48 (0.35-0.60) | 0.52 (0.39-0.65) | 0.86 (0.75-0.93) | 0.98 (0.91-1) |
| PPV | 0.24 (0.13-0.39) | 0.52 (0.39–0.65) | 0.62 (0.40–0.81) | 0.80 (0.28-0.99) |
| NPV | 0.47 (0.35-0.60) | 0.76 (0.60-0.87) | 0.65 (0.54–0.75) | 0.61 (0.51-0.70) |
| OR | 0.29 (0.13-0.67) | 3.39 (1.48-7.78) | 3.11 (1.23-7.80) | 6.24 (0.67-58.4) |

B

| Score | Steroid Non-response |
|---|---|
| 0 | 11/45 (24.4%) |
| 1 | 19/41 (46.3%) |
| 2 | 11/19 (57.9%) |
| 3 | 4/5 (80%) |

**Supplemental Table 7** *Summary statistics for final model performance predicting steroid response using ADMIT-ASC scores of 0-2 as thresholds in the discovery, validation, and combined cohorts. Prop – proportion of patients at or below threshold, sens – sensitivity, spec – specificity, PPV/NPV – positive/negative predictive value, OR – odds ratio. All statistics show 95% confidence intervals in parentheses.*

| | Discovery (Oxford) | | | Validation (Gold Coast) | | | Validation (India) | | | Combined validation | | |
|---|---|---|---|---|---|---|---|---|---|---|---|---|
| Score | 0 | ≤1 | ≤2 | 0 | ≤1 | ≤2 | 0 | ≤1 | ≤2 | 0 | ≤1 | ≤2 |
| Prop | 2.3% | 48.1% | 78.6% | 6.4% | 54.5% | 81.8% | 1.6% | 58.1% | 91.9% | 4.1% | 49.7% | 76.2% |
| Sens | 0.058 (0.00-0.12) | 0.77 (0.65-0.88) | 0.94 (0.88-1.00) | 0.11 (0.03-0.18) | 0.71 (0.60-0.82) | 0.95 (0.90-1.00) | 0.02 (0.00-0.07) | 0.68 (0.54-0.83) | 0.98 (0.93-1.00) | 0.08 (0.03-0.13) | 0.70 (0.61-0.79) | 0.96 (0.93-1.00) |
| Spec | 1.00 (1.00-1.00) | 0.71 (0.61-0.81) | 0.32 (0.21-0.42) | 1.00 (1.00-1.00) | 0.69 (0.55-0.82) | 0.38 (0.24-0.52) | 1.00 (1.00-1.00) | 0.62 (0.41-0.83) | 0.19 (0.02-0.36) | 1.00 (1.00-1.00) | 0.67 (0.55-0.78) | 0.32 (0.21-0.43) |
| PPV | 1.00 (1.00-1.00) | 0.63 (0.52-0.75) | 0.48 (0.38-0.57) | 1.00 (1.00-1.00) | 0.77 (0.66-0.87) | 0.69 (0.59-0.78) | 1.00 (1.00-1.00) | 0.78 (0.64-0.91) | 0.70 (0.58-0.82) | 1.00 (1.00-1.00) | 0.77 (0.69-0.85) | 0.69 (0.62-0.77) |
| NPV | 0.62 (0.53-0.70) | 0.82 (0.73-0.91) | 0.89 (0.78-1.00) | 0.44 (0.34-0.53) | 0.62 (0.49-0.75) | 0.85 (0.69-1.00) | 0.34 (0.23-0.46) | 0.50 (0.31-0.69) | 0.80 (0.45-1.00) | 0.40 (0.33-0.48) | 0.58 (0.47-0.69) | 0.84 (0.70-0.98) |
| OR | Inf | 8.1 (7.3-8.9) | 7.6 (6.3-8.8) | Inf | 5.4 (4.5-6.2) | 13.0 (11.0-14.0) | Inf | 3.5 (2.4-4.6) | 94 (7.1-11.1) | Inf | 4.6 (4.0-5.3) | 11.9 (10.8-13.0) |

**Supplemental Table 8** Summary statistics from validation with the complete Gold Coast validation cohort as shown in Table 6 (left) and results from bootstrapped validation with the same cohort (right, median value with 2.5-97.5 percentile ranges, 1000 rounds, 110 samples).

| | Validation cohort | Bootstrapped results |
|---|---|---|
| Proportion scoring 3+ | 18.2% | 18.1% (10.9-25.5%) |
| Sensitivity | 0.38 (0.24-0.52) | 0.38 (0.24-0.53) |
| Specificity | 0.95 (0.90-1.00) | 0.95 (0.90-1.00) |
| Positive predictive value | 0.85 (0.69-1.00) | 0.85 (0.67-1.00) |
| Negative predictive value | 0.69 (0.59-0.78) | 0.69 (0.59-0.78) |
| Odds ratio | 12.5 (11.2-13.9) | 13.0 (4.2-Inf) |
| Number needed to screen | 6.5 (4.5-11.5) | 6.5 (4.6-11.0) |

**Supplemental Table 9:** *Admission demographics for the Gold Coast cohort. P values for steroid response vs non-response groups from Wilcoxon rank sum test, or Fisher's exact test where marked* [F].

| | Steroid non- response (n=45) | Steroid response (n=65) | p value |
|---|---|---|---|
| Gender | | | 0.85[F] |
| Male (n, %) | 23 (51%) | 31 (47.7%) | |
| Female (n, %) | 22 (49%) | 34 (52.3%) | |
| Median age (range), years | 33 (24-46) | 37 (25-58) | 0.43 |
| Age ≥ 60 years (n, %) | 7 (15.5%) | 15 (23%) | 0.47[F] |
| Median age at diagnosis (range), years | 27 (22-43) | 31 (23-46) | 0.50 |
| Median disease duration (IQR), years | 0.5 (0-4) | 3 (0.2-8) | 0.04 |
| Index presentation as ASC (n, %) | 18 (40%) | 17 (26%) | 0.13 |
| Median follow up post ASC (range), weeks | 100<br>(30-192) | 65<br>(32-168) | 0.50 |
| EIM (n, %) | 4(9%) | 9 (13.8%) | 0.43 |
| 5-ASA (n, %) | | | |
| Current | 22 (48.9%) | 31(47.7%) | 1.00[F] |
| Never | 16 (35.6%) | 19(29.2%) | 0.54[F] |
| Intolerant/ceased | 7 (15.5%) | 15(23.1%) | 0.47[F] |
| IM: methotrexate or thiopurine (n, %) | | | |
| Current | 6 (13.3%) | 6 (9.2%) | 0.54[F] |
| Never | 34 (7.6%) | 46 (70.8%) | 0.67[F] |
| Intolerant/ceased | 5 (11.1%) | 13 (20%) | 0.29[F] |
| Anti-TNF (n, %) | | | |
| Current | 4 (8.9%) | 4 (6.1%) | 0.71[F] |
| Never | 39 (86.7%) | 56 (86.1%) | 1.00[F] |
| Intolerant | 1 (2.2%) | 3 (4.6%) | 0.64[F] |
| Primary non-response | 0 (0%) | 0 (0%) | |
| Secondary loss of response | 1 (2.2%) | 2 (3%) | 1.00[F] |
| Vedolizumab (n, %) | | | |
| Current | 1 (2.2%) | 3 (4.6%) | 0.64[F] |
| Never | 44(97.8%) | 61 (93.9%) | 0.65[F] |
| Intolerant | 0 (0%) | 0 (0%) | - |
| Primary non-response | 0 (0%) | 0 (0%) | - |
| Secondary loss of response | 0 (0%) | 1 (1.5%) | 1.00[F] |
| Tofacitinib | | | |
| Current | 0 (0%) | 0 (0%) | - |
| Never | 0 (0%) | 0 (0%) | - |
| Intolerant | 0 (0%) | 0 (0%) | - |
| Primary non-response | 0 (0%) | 0 (0%) | - |
| Secondary loss of response | 0 (0%) | 0 (0%) | - |
| Previous clinical trial exposure | 0 (0%) | 0 (0%) | - |
| More than 1 advanced therapy exposure (n, %) | 1 (2.2%) | 2 (3%) | 0.82[F] |

**Supplemental Table 10:** *Day 1 admission clinical, laboratory and endoscopic data for Gold Coast Data. P values from Wilcoxon rank sum test, of Fisher's exact test where marked* [F]

| | **Steroid non responder (n=45)** | **Steroid responder (n=65)** | **p value** |
|---|---|---|---|
| Tachycardia >90 bpm (n. %) | 33 (73%) | 28 (43%) | 0.002 [F] |
| Median stool frequency (IQR) | 10 (9-15) | 10 (8-20) | 0.65 |
| Median haemoglobin (IQR), g/L | 121 (102-137) | 125 (111-140) | 0.29 |
| Median CRP (IQR), mg/L | 98 (48-132) | 45 (22-88) | 0.005 |
| Median albumin (IQR), g/L | 29 (25-35) | 35 (31-38) | 0.0001 |
| Median platelet count (IQR), x$10^9$/l | 372 (291-533) | 361 (289-493) | 0.79 |
| Toxic megacolon (n, %) | 1 (2%) | 0 | 0.41 [F] |
| Truelove and Witts' criteria (n, %) | | | *0.015* [F] |
| 1 | 13 (29%) | 37 (57%) | 0.006 [F] |
| 2 | 12 (27%) | 15 (23%) | 0.82 [F] |
| 3 | 16 (36%) | 11 (17%) | 0.04 [F] |
| 4 | 4 (9%) | 2 (3%) | 0.22 [F] |
| UCEIS score (n, %) | | | *0.008* [F] |
| 0-2 | 0 | 3 (5%) | 0.27 [F] |
| 3 | 0 | 4 (6%) | 0.14 [F] |
| 4 | 4 (9%) | 12 (18%) | 0.18 [F] |
| 5 | 15 (33%) | 25 (38%) | 0.69 [F] |
| 6 | 10 (22%) | 15 (23%) | 1 [F] |
| 7 | 13(29%) | 6 (9%) | 0.01 [F] |
| 8 | 3 (7%) | 0 (0) | 0.07 [F] |

**Supplemental Table 11:** *Univariable logistic regression results in the 3 cohorts. Shown as FDR corrected p value (OR, 95% confidence interval).*

| | Oxford | | Gold Coast | AIIMS | |
|---|---|---|---|---|---|
| | Steroid Non-response | Rescue Therapy | Rescue Therapy | Steroid Non-response | Rescue Therapy |
| Age | 0.86 (1.00, 0.98-1.02) | 0.26 (1.00, 0.98-1.02) | | 0.11 (1.04, 0.99 - 1.09) | 0.17 (1.03, 0.99 - 1.08) |
| Sex (Male) | 0.86 (1.12, 0.55-2.29) | 0.47 (1.50, 0.75-3.05) | 0.80 (1.10, 0.54-2.22) | 0.27 (0.46, 0.14 - 1.39) | 0.15 (0.35, 0.09 - 1.17) |
| Disease Duration | 0.62 (1.01, 0.98-1.05) | 0.81 (0.99, 0.96-1.03) | 0.70 (0.99, 0.98-1.01) | 0.22 (1.09, 0.97 - 1.31) | 0.15 (1.12, 0.99 - 1.36) |
| First Admission | 0.70 (0.58, 0.15-1.83) | 0.82 (1.21, 0.39-3.74) | 0.17 (1.64, 0.77-3.48) | | |
| Current biologic | 0.82 (1.21, 0.57-2.63) | 0.48 (1.49, 0.71-3.21) | | | |
| Albumin | **0.0066 (0.89, 0.83-0.95)** | **0.029 (0.91, 0.85-0.97)** | **<0.001 (0.88, 0.83-0.94)** | 0.51 (0.97, 0.90 - 1.05) | 0.97 (1.00, 0.92 - 1.09) |
| CRP | **0.00066 (1.02, 1.01-1.03)** | **0.0091 (1.01, 1.01-1.02)** | **0.005 (1.01, 1.01-1.01)** | 0.36 (1.01, 0.99 - 1.02) | 0.84 (1.00, 0.99 - 1.01) |
| Haemoglobin | 0.60 (0.99, 0.98-1.01) | 0.76 (1.00, 0.98-1.03) | 0.10 (0.99, 0.97-1.00) | 0.58 (1.01, 0.98 - 1.04) | 0.94 (1.00, 0.97 - 1.03) |
| Platelets | 0.55 (1.00, 1.00-1.00) | 0.47 (1.00, 1.00-1.01) | 0.91 (0.99-1.00) | 0.36 (1.00, 0.99 - 1.00) | 0.73 (1.00, 0.99 - 1.00) |
| Stool frequency | 0.53 (1.05, 0.97-1.05) | 0.31 (1.08, 0.99-1.18) | 0.66 (0.99, 0.93-1.04) | 0.29 (1.11, 0.91 - 1.36) | 0.21 (1.14, 0.93 - 1.41) |
| TW score | **0.0066 (2.43, 1.45-4.29)** | 0.094 (1.78, 1.10-2.97) | **0.01 (1.64, 1.12-2.39)** | 0.61 (1.17, 0.64 - 2.14) | 0.17 (1.58, 0.84 - 3.10) |
| TW without CRP | 0.19 (1.75, 0.98-3.23) | 0.46 (1.43, 0.82-2.55) | | 0.95 (1.03, 0.46 - 2.29) | 0.34 (1.51, 0.65 - 3.72) |
| UCEIS score | **0.015 (1.62, 1.18-2.27)** | **0.029 (1.57, 1.15-2.18)** | **<0.001 (2.03, 1.15-2.07)** | **0.0008 (3.10, 1.68 - 6.41)** | **0.01 (2.43, 1.35 - 4.85)** |
| UCEIS erosion & ulceration | **0.0066 (2.68, 1.53-5.00)** | **0.029 (2.13, 1.34-4.17)** | | | |

**Supplemental Table 12:** *Admission demographics for the AIIMS cohort. P values for steroid response vs non-response groups from Wilcoxon rank sum test, or Fisher's exact test where marked [F].*

| | Steroid non-response (n=21) | Steroid response (n=41) | p value |
|---|---|---|---|
| Gender<br>Male (n, %)<br>Female (n, %) | <br>6 (28.6%)<br>15 (71.4%) | <br>19 (46.3%)<br>22 (53.7%) | 0.27[F] |
| Median age (range), years | 39 (18-63) | 33 (15-70) | 0.09 |
| Age ≥ 60 years (n, %) | 2 (9.5%) | 1 (2.4%) | 0.26[F] |
| Median age at diagnosis (range), years | 35.9 (17.2-58) | 28.0 (13-69.3) | 0.09 |
| Median disease duration (IQR), years | 2.5 (0.1-32) | 3.0 (0.25-12) | 0.93 |
| EIM (n, %) | 8 (38.1%) | 8 (19.5%) | 0.13 |

**Supplemental Table 13:** *Day 1 admission clinical, laboratory and endoscopic data for AIIMS. P values from Wilcoxon rank sum test, of Fisher's exact test where marked [F]*

| | **Steroid non responder (n=21)** | **Steroid responder (n=41)** | **p value** |
|---|---|---|---|
| Tachycardia >90 bpm (n. %) | 17 (81%) | 34 (83%) | 1.00[F] |
| Median stool frequency (IQR) | 10 (9-12) | 9 (8-10) | 0.09 |
| Median haemoglobin (IQR), g/L | 94 (87-112) | 96 (81-109) | 0.60 |
| Median CRP (IQR), mg/L | 25 (7-46) | 24 (9-35) | 0.85 |
| Median albumin (IQR), g/L | 30 (26-34) | 32 (27-35) | 0.41 |
| Median platelet count (IQR), $x10^9/l$ | 288 (250-310) | 310 (212-372) | 0.49 |
| Toxic megacolon (n, %) | 0 | 0 | - |
| Truelove and Witts' criteria (n, %)<br>1<br>2<br>3<br>4 | <br>9 (43%)<br>3 (14%)<br>9 (43%)<br>0 (0%) | <br>15 (37%)<br>16 (39%)<br>9 (22%)<br>1 (2%) | 0.13[F]<br>0.78[F]<br>0.08[F]<br>0.14[F]<br>1.00[F] |
| UCEIS score (n, %)<br>3<br>4<br>5<br>6<br>7<br>8 | <br>1 (5%)<br>0 (0%)<br>2 (10%)<br>8 (38%)<br>9(43%)<br>1 (5%) | <br>1 (2%)<br>6 (15%)<br>22 (54%)<br>7 (18%)<br>5 (12%)<br>0 (0%) | *0.0001*[F]<br>1.00[F]<br>0.09[F]<br>0.0008[F]<br>0.12[F]<br>0.01[F]<br>0.34[F] |

**Supplemental Figure 1** *Day 1 CRP, albumin, Truelove & Witts' criteria, and UCEIS scores in steroid response and non-response groups (see Table 2).*

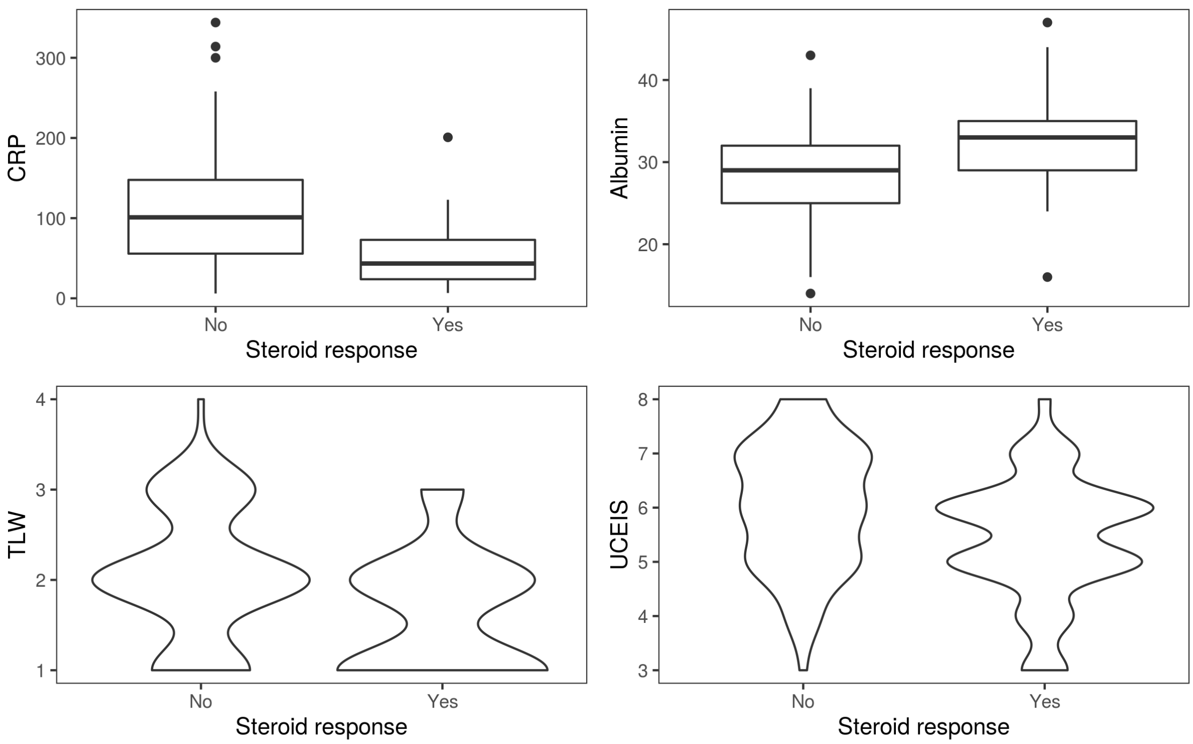

**Supplemental Figure 2:** *Confusion matrices showing classification averages in left out samples from 10 repeats of 10-fold cross-validation optimising for accuracy for the models shown in Supplemental Table 4.*

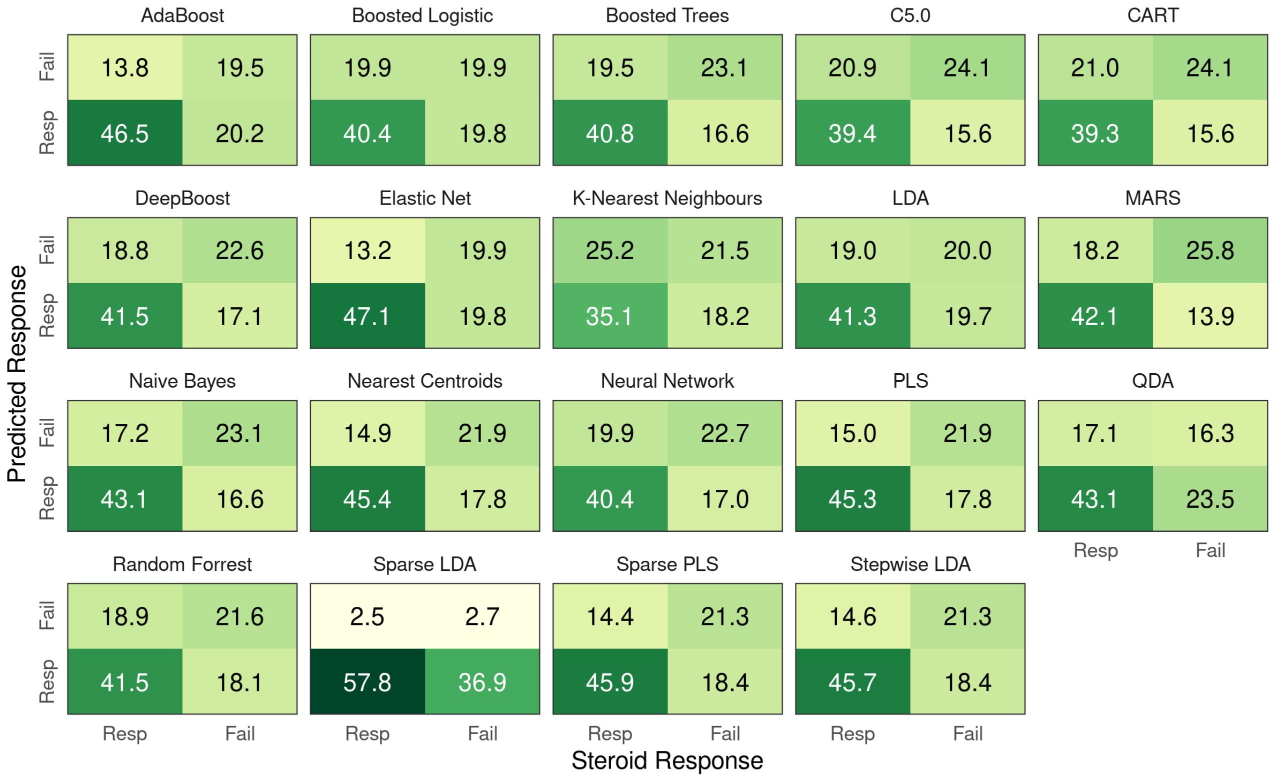

**Supplemental figure 3:** *Variable importance estimates for models shown in supplemental table 4. Models not listed did not have a unique function for assessing variable importance and share results with Boosted Logistic Regression.*

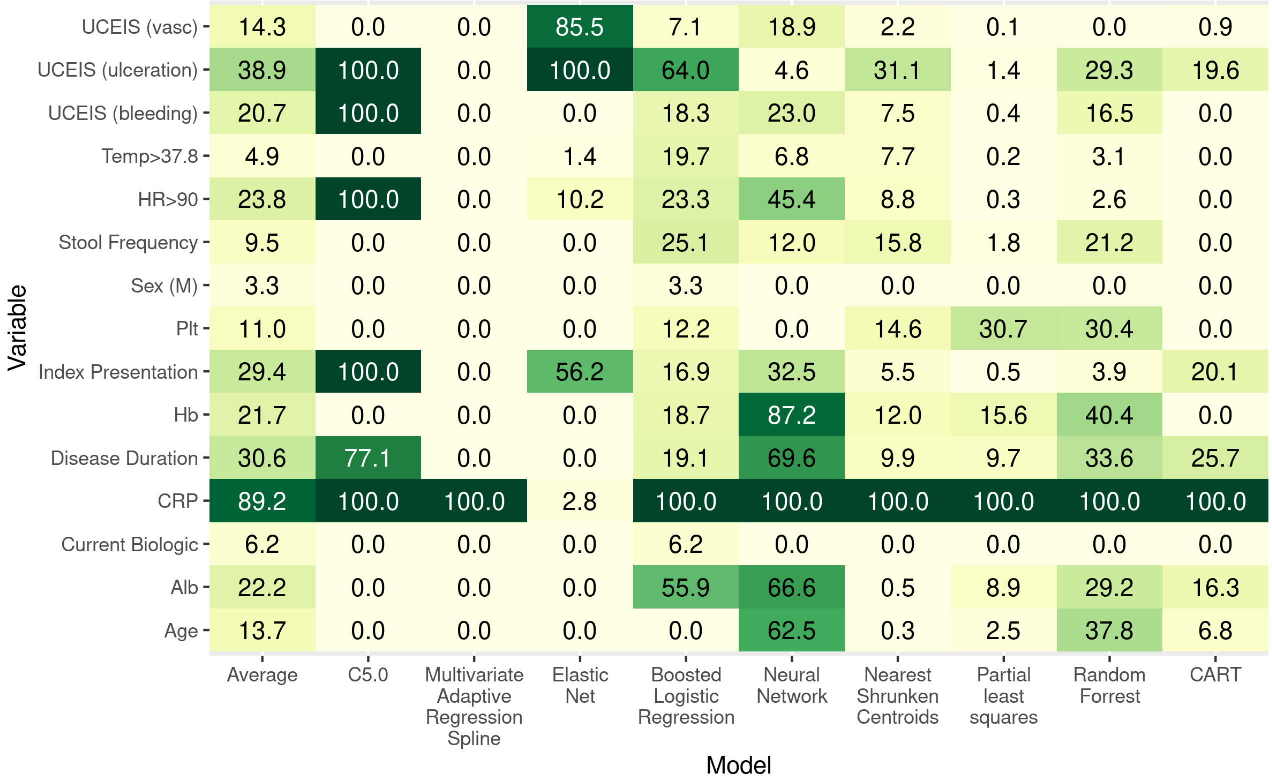

**Supplemental Figure 4:** *ROC curves showing the density of ROC curves in left out samples from each of 10 repeates of 10-fold cross-validation for each model. The blue line shows a LOESS estimate of the average ROC curve and the mean AUC value is shown in the top left.*

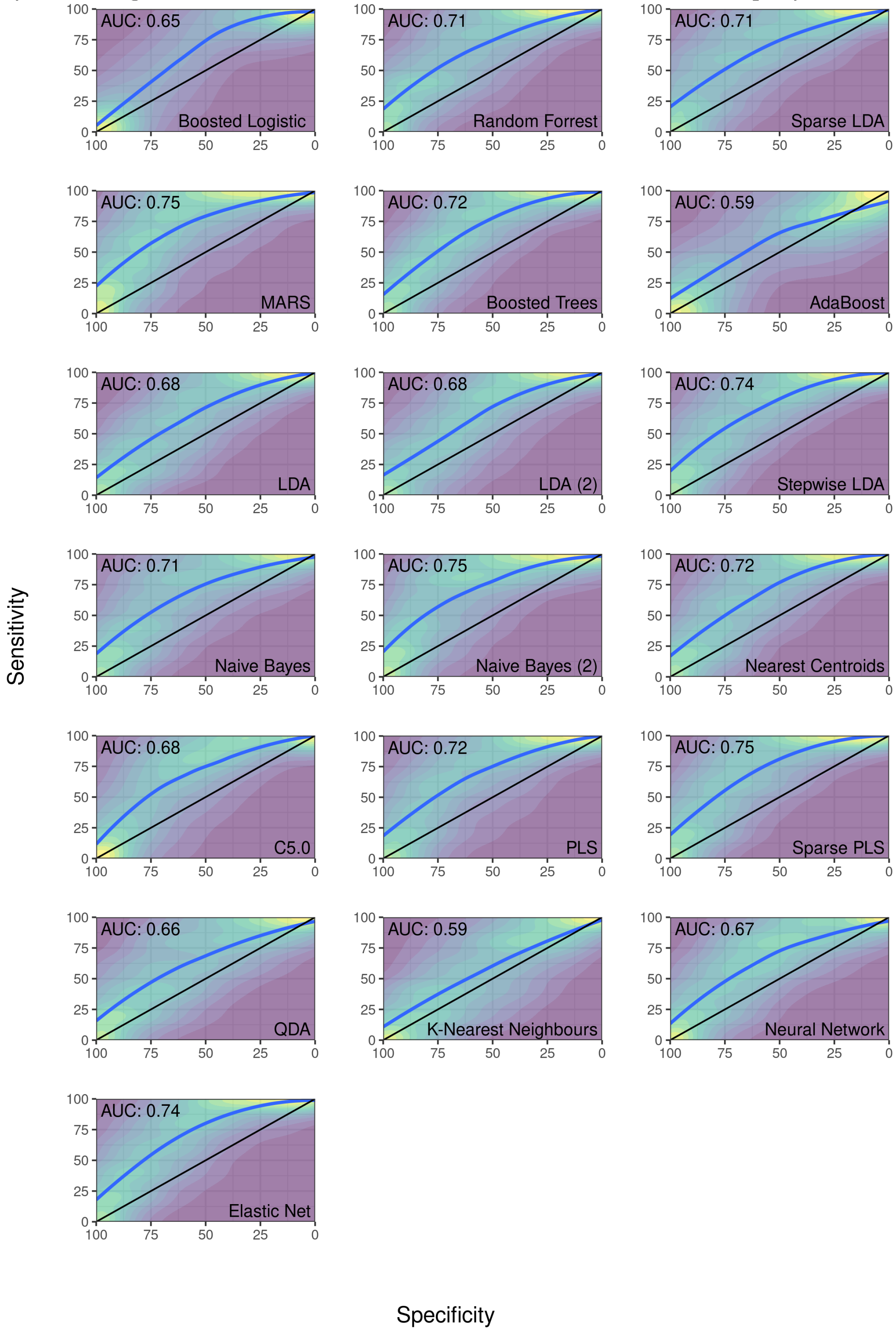
